# Large Language Models Facilitate the Generation of Electronic Health Record Phenotyping Algorithms

**DOI:** 10.1101/2023.12.19.23300230

**Authors:** Chao Yan, Henry H. Ong, Monika E. Grabowska, Matthew S. Krantz, Wu-Chen Su, Alyson L. Dickson, Josh F. Peterson, QiPing Feng, Dan M. Roden, C. Michael Stein, V. Eric Kerchberger, Bradley A. Malin, Wei-Qi Wei

## Abstract

**Objectives:** Phenotyping is a core task in observational health research utilizing electronic health records (EHRs). Developing an accurate algorithm demands substantial input from domain experts, involving extensive literature review and evidence synthesis. This burdensome process limits scalability and delays knowledge discovery. We investigate the potential for leveraging large language models (LLMs) to enhance the efficiency of EHR phenotyping by generating high-quality algorithm drafts.

**Materials and Methods:** We prompted four LLMs—GPT-4 and GPT-3.5 of ChatGPT, Claude 2, and Bard—in October 2023, asking them to generate executable phenotyping algorithms in the form of SQL queries adhering to a common data model (CDM) for three phenotypes (i.e., type 2 diabetes mellitus, dementia, and hypothyroidism). Three phenotyping experts evaluated the returned algorithms across several critical metrics. We further implemented the top-rated algorithms and compared them against clinician-validated phenotyping algorithms from the Electronic Medical Records and Genomics (eMERGE) network.

**Results:** GPT-4 and GPT-3.5 exhibited significantly higher overall expert evaluation scores in instruction following, algorithmic logic, and SQL executability, when compared to Claude 2 and Bard. Although GPT-4 and GPT-3.5 effectively identified relevant clinical concepts, they exhibited immature capability in organizing phenotyping criteria with the proper logic, leading to phenotyping algorithms that were either excessively restrictive (with low recall) or overly broad (with low positive predictive values).

**Conclusion:** GPT versions 3.5 and 4 are capable of drafting phenotyping algorithms by identifying relevant clinical criteria aligned with a CDM. However, expertise in informatics and clinical experience is still required to assess and further refine generated algorithms.

## INTRODUCTION

Electronic health record (EHR) phenotyping, which involves creating algorithms to identify and correctly classify a patient’s observable characteristics by integrating complex clinical data, has become pivotal in observational health research^1^. Developing EHR phenotypes is an intricate and labor-intensive process that demands extensive expertise in both the clinical and informatics domains^2,3^. While phenotyping includes the identification of individuals with specific characteristics, it also necessitates the selection of suitable controls for meaningful comparisons with the identified cases^4^.

Rule-based computable phenotyping algorithms rely on clinical experts to select specialized criteria (e.g., diagnosis codes, medications, and laboratory values) likely to define a phenotype of interest. When subjected to detailed refinement and thorough validation, these algorithms often exhibit enhanced performance compared to high-throughput methods, which typically employ machine learning or data mining-based approaches to provide automated and rapid categorization of numerous disease phenotypes.^5–8^ However, the iterative nature of this process often requires substantial literature review and discussions with clinical experts to generate a single phenotyping algorithm, thereby limiting the scalability of this approach in practice^2^.

Furthermore, implementation of phenotyping algorithms by secondary sites requires additional informatics expertise, manual effort, and time to adapt the existing code to different databases and EHR systems.

Recently, large language models (LLMs) have demonstrated effectiveness in information extraction and summarization^9^, indicating a potential benefit in phenotyping by reducing the time required for literature review and synthesis during the phenotype generation process. Previous studies investigating the application of LLMs to phenotyping tasks have primarily evaluated the ability of LLMs to extract phenotypic information from unstructured clinical notes^10^. For example, Alsentzer et al. found that the open source LLM Flan-T5 could effectively extract concepts from discharge summaries to create a postpartum hemorrhage phenotype^11^. In this preliminary report, we investigate the novel application of LLMs for generating computable phenotyping algorithms to assess whether such tools can effectively expedite the development of EHR phenotypes based on structured data. We appraised four LLMs—GPT-4 (powering ChatGPT)^12^, GPT-3.5 (powering ChatGPT)^13^, Claude 2^14^, and PaLM 2-powered Bard^15^—to generate phenotyping algorithms for three clinical phenotypes—type 2 diabetes mellitus (T2DM), dementia, and hypothyroidism. We subsequently implemented the top-rated algorithms as adjudicated by phenotyping experts using multiple critical metrics from each LLM and compared them against the clinician-validated phenotyping algorithms from the Electronic Medical Records and Genomics (eMERGE) network^16,17^.

## METHODS

We selected four LLMs in their default configurations to test their phenotyping algorithm generation capacity for three common clinical phenotypes. These LLMs were: 1) GPT-4 and 2) GPT-3.5 (both powering ChatGPT by OpenAI)^12,13^, 3) Claude 2 (developed by Anthropic)^14^, and 4) Bard (created by Google and based on PaLM 2)^15^. These models were chosen because of their widespread use, easy accessibility, extensive evaluation, robust computational capabilities and proficiency in handling and generating lengthy texts—qualities crucial for sustainably supporting phenotyping tasks.

This pilot study specifically focused on three clinical phenotypes: T2DM^18,19^, dementia^20^, and hypothyroidism^21,22^. We chose these phenotypes because they have existing algorithms that have undergone extensive validation processes and demonstrated highly accurate and consistent performances with well-documented results. Data collection via web-based interactions with the LLMs occurred in October 2023, with subsequent data analysis completed in November 2023. This study was approved by the institutional review boards at Vanderbilt University Medical Center (IRB #: 201434).

**Figure 1** illustrates an overview of the study pipeline, which was comprised of two main components—prompting (steps 1-3, highlighted in pink) and evaluating (steps 4-9, highlighted in blue) LLMs.

**Figure 1.**
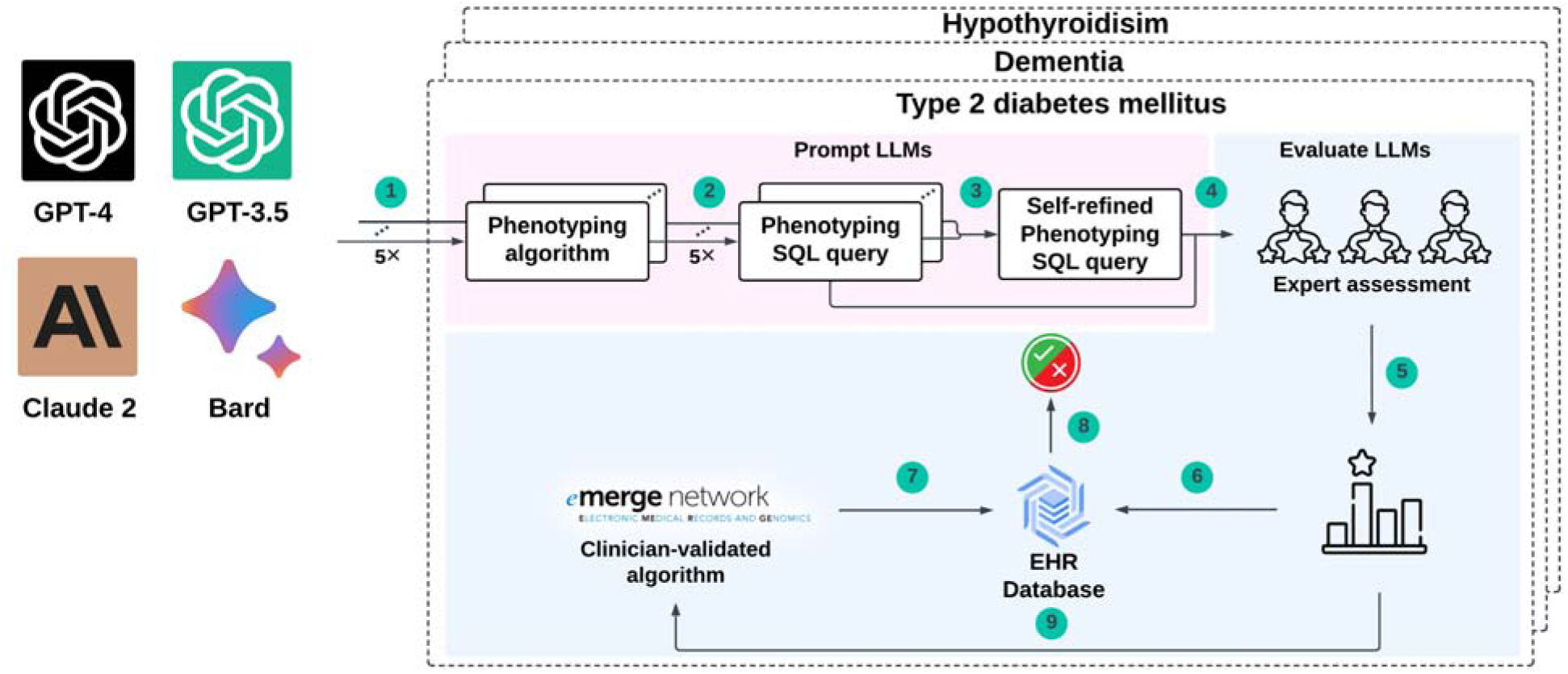
An architectural overview of the study pipeline.

### Prompting large language models to generate phenotyping algorithms

We prompted the LLMs to generate executable SQL queries to identify phenotype cases from structured EHR data organized according to the Observational Medical Outcomes Partnership (OMOP) Common Data Model (CDM), a standard EHR data framework that enables efficient data analysis and sharing across institutions^23,24^. There are multiple methods to search for concepts in an OMOP database, including specifying “concept_code”, “concept_name”, or “concept_id”. In this study, we focused on using ICD codes as concept codes for diagnosis concepts and using concept names for non-diagnosis concepts. This design was based on our observations that LLMs were 1) able to generate ICD diagnosis codes that are relevant to the target phenotype; 2) less likely to identify meaningful non-diagnosis concepts using “concept_code” compared to using “concept_name”; and 3) unable to generate applicable“concept_id” in general. The variability in the capacity of LLMs to identify relevant concepts through different methods can be attributed to discrepancies in the amount of information they have encountered about these methods to during pretraining.

We designed two distinct prompting strategies, hereafter termed α-prompting and β-prompting. The α-prompting strategy (steps 1 and 2 in **Figure 1**) had two steps. The first step focused on obtaining a pseudocode version of the phenotyping algorithm (referred to as a pseudo-phenotyping algorithm), which emphasized identifying and integrating critical phenotyping criteria, as well as determining the strategy for combining these criteria. We utilized the chain-of-thought (CoT) prompting strategy^25^, an effective method for directing LLMs through a series of reasoning steps to resolve complex problems like humans. Specifically, we framed our instruction to guide reasoning as follows: “Let’s think step by step: 1. List the critical criteria to consider. 2. Determine how these criteria should be combined. 3. Derive the final algorithm” (Supplemental **Table 1**). Additionally, multiple detailed instructions were specified so that the produced pseudo-phenotyping algorithm adhered to the OMOP concepts, including diagnosis codes (in ICD-9-CM and ICD-10-CM), symptoms, procedures, laboratory tests, and medications (both generic and brand names). We also mandated that the pseudo-phenotyping algorithm maintain a style consistent with the SQL logic, to facilitate generation of the SQL query in the second step.

Using the response of an LLM in the initial step, the second step involved converting the pseudo-phenotyping algorithm into an executable SQL query (referred to as an SQL-formatted phenotyping algorithm) for implementation in an OMOP CDM-based EHR database for subsequent validation (Supplemental **Table 1**). Due to the probabilistic nature of LLMs, variations in the generated phenotyping algorithms are guaranteed. Consequently, we executed α-prompting five times independently for each phenotype and for each LLM to account for response variability.

The β-prompting strategy (step 3 in **Figure 1**) was designed to first present the LLM with the five SQL-formatted phenotyping algorithms generated from the α-prompting strategy and then instruct the LLM to assess the quality of these algorithms and generate an improved one (Supplemental **Table 1**). This strategy, proven to effectively mitigate hallucinations^26–28^ (i.e., in the context of EHR phenotyping, generating column names that do not follow OMOP CDM or producing concept names, concept IDs, or even logics that are irrelevant to the target phenotype), leverages an LLM’s ability to evaluate scientific texts based on the extensive knowledge encoded during model pre-training. As a result, the LLM can produce an updated version of the phenotyping algorithm through analytical reflections. We also deployed the CoT strategy in β-prompting, which involved initially identifying the correct, incorrect, and missing criteria for each previously generated algorithm. The β-prompting strategy was executed in an independent session of the LLM (distinct from and subsequent to those used for α-prompting) and was executed only once.

### Evaluating the quality of LLM-generated phenotyping algorithms

We then performed a comprehensive analysis, encompassing both qualitative and quantitative assessments, to evaluate the efficacy of the phenotyping algorithms generated by the four different LLMs on the three diseases of focus as described above. For the qualitative analysis, three experts (W.Q.W., M.E.G., and V.E.K.) in EHR phenotyping and clinical medicine, each with significant experience in clinical and informatics research, independently reviewed and rated the LLM-generated SQL-formatted phenotyping algorithms in a blind manner. All experts had authored numerous papers related to clinical phenotyping. We further compared the concepts utilized in these algorithms with those found in clinician-validated phenotyping algorithms developed by the eMERGE network^16,17^. The algorithms selected^18–22^ were developed among large patient populations, validated across multiple research centers, and widely recognized for their reliability. Specifically for dementia, the chosen eMERGE algorithm excludes mild cognitive impairment codes and does not filter out delirium diagnoses, reflecting the reality that dementia patients may also present with delirium. For quantitative assessment, we implemented the top-rated SQL-formatted phenotyping algorithms using EHR data from Vanderbilt University Medical Center (VUMC) and assessed their performance against established eMERGE algorithms^18–22^.

### Expert assessment

Each of the three experts conducted independent reviews for every SQL-formatted phenotyping algorithm (4 LLMs, 3 phenotypes, and 2 strategies, for 24 algorithms in total). The evaluation focused on three dimensions: 1) adherence to instructions, which assessed how well the LLM conformed to predefined formatting rules; 2) the generation of proficient phenotyping algorithms based on knowledge, which evaluated the LLM’s ability to synthesize and organize phenotyping-related information effectively; and 3) presentation in executable SQL format, which measured the potential of an LLM to reduce the labor-intensive human efforts required for EHR implementation and validation. Detailed guidelines for expert evaluation can be found in Supplemental **Table 2**. Experts assigned categorical scores (“Good [3],” “Medium [2],” or “Poor [1]”) for each axis based on predefined criteria, providing justifications accordingly. Interrater reliability was assessed using the weighted Cohen’s Kappa score^29^. We compared LLMs’ rated scores using the Wilcoxon signed-rank test^30^ with a significance level of 0.05.

### Comparison of concept coverage with eMERGE phenotyping algorithms

This analysis provided a comprehensive comparison of the concepts within phenotyping algorithms generated by LLMs and established EHR algorithms. We systematically reviewed and compared all the concepts employed in the algorithms for diagnoses, laboratory tests, procedures, medications, symptoms, and exclusions, and then summarized the noteworthy findings.

### Implementation of LLM-generated algorithms and performance evaluation

We deployed the highest-rated phenotyping algorithms for each phenotype in a research cohort at VUMC (n=84,821). This cohort, extensively utilized in phenotyping research, has been a significant resource for phenotypic studies^5,31^. We summarized implementation details in Supplemental Section S.3, where we list the general edits we made as well as specific changes for each implemented algorithm (Supplemental **Table 3**). As a benchmark, we implemented three eMERGE algorithms updated with current ICD-10-CM codes^5^ to identify phenotype cases and controls. The cases and controls identified by the eMERGE algorithms served as a reference standard to assess the effectiveness of the LLM-generated algorithms. In our subsequent analysis, we excluded patients not categorized as either case or control by the eMERGE algorithm, as their data did not meet the criteria for either situation. Each of the top-rated LLM-generated algorithms required some modifications to be executable in a cloud-based platform to securely query VUMC’s research clinical databases that follow the OMOP CDM. We limited changes to technical domain knowledge, as opposed to clinical domain knowledge. For example, we edited the database names, but did not edit drug names or ICD codes.

We used the following metrics for evaluation: 1) positive predictive values (PPV), defined as the number of cases mutually identified by the eMERGE algorithm and an LLM over the total number of identified cases by the LLM; 2) recall, defined as the number of cases mutually identified by the eMERGE algorithm and an LLM over the total number of cases identified by the eMERGE algorithm; and 3) false positive rate (FPR), defined as the number of patients identified as cases by an LLM but identified as controls by the eMERGE algorithm (false positives) over the sum of false positives and number of cases identified by the eMERGE algorithm.

## RESULTS

### Expert assessments

The average interrater reliability was 0.59 [0.50-0.68], indicating a moderate to substantial agreement among experts, considering the categorical nature of the data (rather than dichotomous) and the variations in scoring criteria among different experts^32^.

By mapping the experts’ assessments of “Good”, “Medium”, or “Poor” to numerical scores of 3, 2, and 1, respectively, GPT-4 (mean [95% confidence interval]: 2.57 [2.40-2.75]) and GPT-3.5 (2.43 [2.25-2.60]) exhibited significantly higher overall expert evaluation scores than Claude 2 (1.91 [1.68-2.13]) and Bard (1.20 [1.09-1.31]) (**Figure 2A**). GPT-4 marginally outperformed GPT-3.5, though the differences were not statistically significant. Moreover, the β-prompting strategy did not significantly differ from the α-prompting, according to experts’ evaluation (**Figure 2B**). Furthermore, experts assigned higher scores to LLMs for their effectiveness in generating phenotyping algorithms for T2DM and hypothyroidism compared to dementia (**Figure 2C**). The radar plot shown in **Figure 2D** displays the average scores for each involved LLM across the three axes of evaluation. There are two key findings. First, GPT-4 and GPT-3.5 were rated consistently rated better than Claude 2 and Bard in following instructions, algorithmic logic, and SQL executability. Second, GPT-4 was considered to be on par with GPT-3.5 in its ability to follow instructions and SQL executability, yet it surpassed GPT-3.5 in its algorithmic logic. Based on these findings, we continued our investigation with GPT-4 and GPT-3.5.

**Figure 2.**
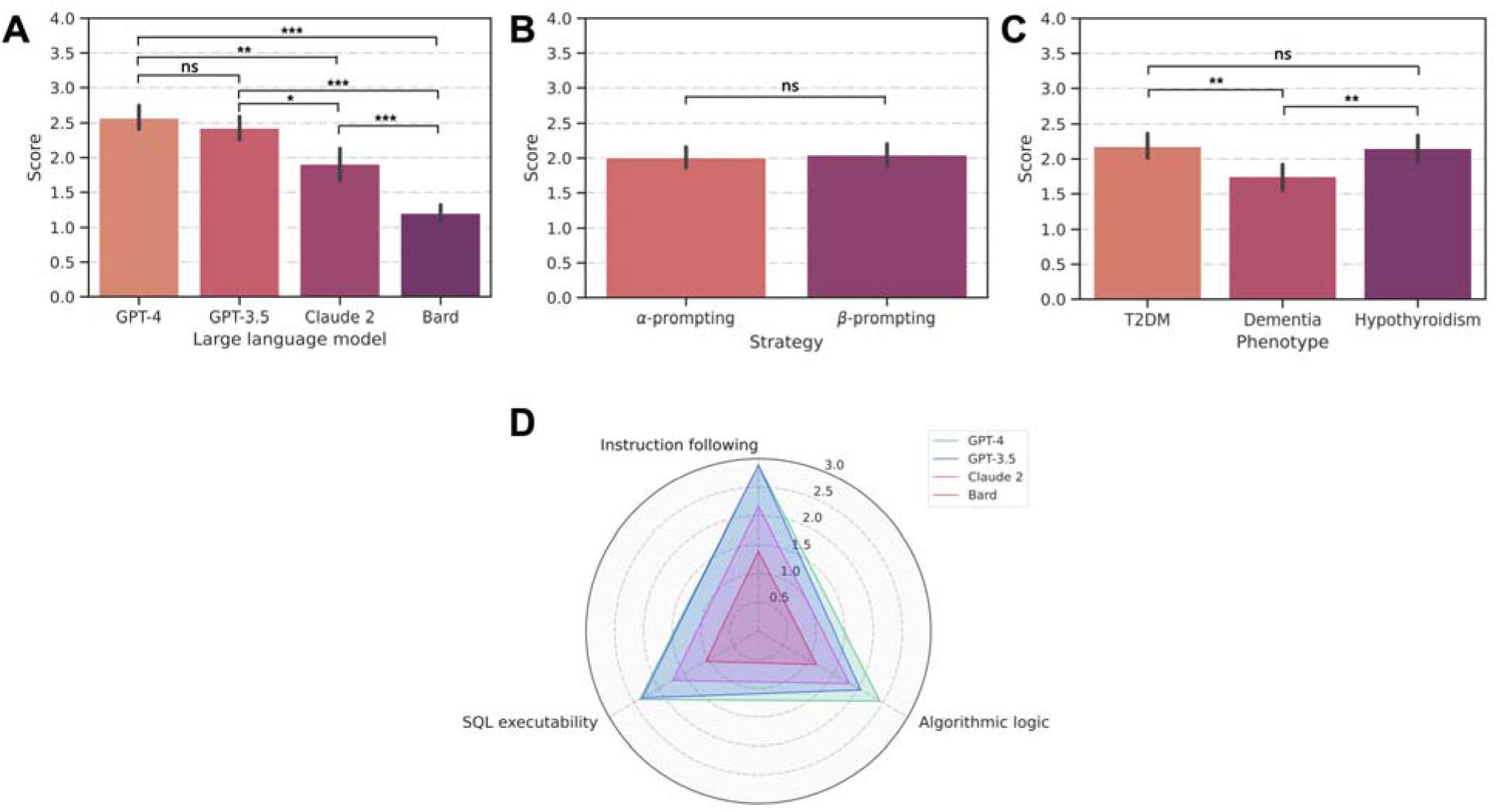
A comparative analysis based on expert evaluations focusing on A) four large language models, B) two prompting strategies, C) three phenotypes, and D) three individual evaluation axes. Numeric scores of 3, 2, and 1 correspond to expert assessments of “Good”, “Medium”, and “Poor”, respectively. ***, **, and * denote *p*<0.001, *p*<0.01, and *p*<0.05, respectively. ns=not significant.

### Comparison with eMERGE phenotyping algorithms

**Components Table 1** summarizes the clinical concepts identified by the eMERGE phenotyping algorithms and LLM-produced algorithms. A full comparison of concepts can be found in Supplemental **Table 4**. Given that the phenotyping algorithms produced by both GPT-4 and GPT-3.5 from the β-prompting strategy were rated similarly to those from the α-prompting strategy, we focused further analyses on the β-prompting results.

**Table 1.** Shared and non-shared concepts from the eMERGE, GPT-4 (β-prompting), and GPT-3.5 (β-prompting) phenotyping algorithms for T2DM, dementia, and hypothyroidism. Darker blue shading represents a relatively higher count of unshared concepts when comparing the algorithms. Orange shading represents inaccurate concepts or laboratory cutoffs.

| Type 2 diabetes mellitus |  |  |  |  |
| --- | --- | --- | --- | --- |
| Concept | Shared | Unshared concept count (example) |  |  |
| Algorithm | All | eMERGE | GPT-4 (β) | GPT-3.5 (β) |
| Diagnoses | 250.*0, 250.*2, E11.* | 2<br>(O24.11) | 0 | 31<sup>a</sup><br>(250.01) |
| Lab tests or procedures | HgbA1c<sup>b</sup>, Fasting BG<sup>c</sup> | 1 Lab<br>(Random BG >200)<sup>d</sup> | 1 Lab<br>(Oral GTT >200)<sup>d</sup> | 1 Lab<br>(BMI ≥25)<sup>d</sup> |
| Medications | Metformin, Glipizide | 34<br>(Exenatide)<sup>d</sup> | 7<br>(Glucophage) | 1<br>(Sitagliptin) |
| Symptoms | None | 0 | 3<br>(Polyuria) | 3<br>(Polydipsia) |
| Exclusion by Type | None | 9 ICD<br>(250.*1) | 3 ICD<br>(250.*3) | 0 |
| Dementia |  |  |  |  |
| Concept | Shared | Unshared concept count |  |  |

|  |  | (example) |  |  |
| --- | --- | --- | --- | --- |
| Algorithm | All | eMERGE | GPT-4 ($\beta$) | GPT-3.5 ($\beta$) |
| Diagnoses | None | 45<br>(290.0)<sup>d</sup> | 6 | 20 |
| Lab tests or procedures | None | 0 | 4 Procedures<br>(MMSE)<sup>d</sup> | 3 Procedures<br>(MRI scan)<sup>d</sup> |
| Medications | Donepezil, Aricept,<br>Memantine, Namenda | 11<br>(Cognex)<sup>d</sup> | 4<br>(Galantamine) | 0 |
| Symptoms | None | 0 | 3<br>(Impaired reasoning)<sup>d</sup> | 5<br>(Cognitive impairment)<sup>d</sup> |
| Exclusion by Type | None | 0 | 3 Labs<br>(B12 level)<sup>d</sup> | 0 |
| Hypothyroidism |  |  |  |  |
| Concept | Shared | Unshared concept count<br>(example) |  |  |
| Algorithm | All | eMERGE | GPT-4 ($\beta$) | GPT-3.5 ($\beta$) |
| Diagnoses | 244.9, E03.8, E03.9 | 18<br>(244)<sup>d</sup> | 7<br>(244.1)<sup>d</sup> | 6<br>(E03.5)<sup>d</sup> |
| Lab tests or procedures | TSH<sup>e</sup> | 4 Labs<br>(Anti-TPO)<sup>d</sup> | 1 Lab<br>(Serum free thyroxine) | 0 |
| Medications | Levothyroxine, Synthroid | 9<br>(Liothyronine)<sup>d</sup> | 15<br>(Amiodarone)<sup>d,f</sup> | 0 |
| Symptoms | None | 0 | 8<br>(Hair loss)<sup>d</sup> | 8<br>(Constipation)<sup>d</sup> |
| Exclusion by Type | None | 18 ICD<br>(193*)<sup>d</sup><br>79 CPT<br>(60240)<sup>d</sup><br>16 Medications<br>(Amiodarone)<sup>d</sup> | 2 Procedures<sup>g</sup><br>(Thyroid surgery) | 0 |

Abbreviations: BG, blood glucose; BMI, body mass index; CPT, Current Procedural Terminology; GTT, glucose tolerance test; HgbA1c, hemoglobin A1c; ICD, International Classification of Diseases; MMSE, Mini-Mental State Exam; MRI, magnetic resonance imaging; TSH, thyroid stimulating hormone; TPO, thyroperoxidase.
<sup>a</sup> Inaccurately includes IC9-CM codes for type 1 diabetes mellitus with 250\*.
<sup>b</sup> All 3 phenotyping algorithms use a HgbA1c cutoff > 6.5
<sup>c</sup> eMERGE and ChatGPT-4 use the correct fasting BG > 126 mg/dL and ChatGPT-3.5 incorrectly uses cutoff > 6.5 mg/dL
<sup>d</sup> This denotes a unique concept example not used in any other phenotyping algorithm
<sup>e</sup> Different TSH cutoffs used by each phenotyping algorithm (eMERGE: > 5, ChatGPT-4: > upper limit of normal; ChatGPT-3.5: >4.5)
<sup>f</sup> Inaccurately includes amiodarone, which can cause hypothyroidism, as a treatment for hypothyroidism
<sup>g</sup> Rather than specifying specific procedure codes, queried the concept table for concepts exactly matching "Thyroid surgery" or "Radioactive iodine treatment" (returned 5 procedure codes including 2 SNOMED, 1 Nebraska Lexicon, 1 HemOnc, and 1 MeSH)

There are several notable observations. For the T2DM phenotyping algorithm, GPT-4 and GPT-3.5 identified relevant diagnosis codes (both ICD-9-CM and ICD-10-CM), lab tests (hemoglobin A1c, fasting blood glucose), and two generic medications (metformin and glipizide) that were also used in the eMERGE phenotyping algorithm. The eMERGE phenotyping algorithm also included medications not identified by either GPT-4 or GPT-3.5 (n=27). Only GPT-4 and GPT-3.5 included symptoms; both eMERGE and GPT-4 provided exclusionary criteria (ICD codes). Notably, the GPT-3.5 model incorrectly included ICD-9-CM codes for type 1 diabetes and applied an incorrect cutoff value for fasting blood glucose (>6.5 mg/dL instead of >125 mg/dL).

For the dementia phenotyping algorithm, GPT-4 and GPT-3.5 included relevant diagnosis codes (ICD-9-CM and ICD-10-CM, as well as one ICD-10). While there were several overlapping diagnosis codes between each pair of phenotyping algorithms, no diagnosis codes were shared across all three algorithms. GPT-4 and GPT-3.5 included symptoms potentially related to dementia while the eMERGE algorithm did not. All three phenotyping algorithms shared four medications (two generic: donepezil and memantine, and two brand names: Aricept and Namenda), although the eMERGE algorithm specified additional medications that did not appear in either LLM-generated algorithm (n=7). Only the GPT-4 phenotyping algorithm included exclusion criteria (e.g., vitamin B12 level).

For the hypothyroidism phenotyping algorithm, GPT-4 and GPT-3.5 identified relevant diagnosis codes (both ICD-9-CM and ICD-10-CM), lab tests (thyroid stimulating hormone), and medications (generic levothyroxine and brand name Synthroid) used in the eMERGE algorithm. Only the eMERGE algorithm used thyroid autoantibodies (e.g., thyroid antiperoxidase). Notably, the GPT-4 algorithm specified the largest number of medications (n=15), including 9 medications that were not included in either the eMERGE algorithm or the GPT-3.5 algorithm. Once again, only GPT-4 and GPT-3.5 included symptoms. The eMERGE algorithm included 113 exclusions, including 18 ICD codes, 79 CPT codes, and 16 medications, while GPT-4 and GPT-3.5 specified only 2 and 0 exclusions, respectively.

### Implementation and evaluation in VUMC

Along with the eMERGE algorithms, we successfully deployed all algorithms generated by GPT-4 and GPT-3.5 from the β-prompting strategy except the T2DM algorithm generated by GPT-3.5 (**Table 2**). The failure of this phenotyping algorithm stemmed from its restrictive logic which accumulated LEFT JOINs across various tables, including the Person, Condition_Occurrence, Measurement, Drug_Exposure, and Observation tables. The algorithm required a LEFT JOIN on a list of people who had a record of symptoms in the Observation table (Polyuria, Polydipsia, Unexplained weight loss) and had a value of zero in the “value_as_concept_id” column. A value of zero did not exist for these symptoms, therefore the algorithm could not identify any individuals who met this criterion for T2DM.

**Table 2.** Performance of the phenotyping algorithms generated by GPT-4 and GPT-3.5 from the β-prompting strategy when applied to VUMC data, as measured against.

| Disease | eMERGE |  | GPT-4 |  |  |  |  | GPT-3.5 |  |  |  |  |
| --- | --- | --- | --- | --- | --- | --- | --- | --- | --- | --- | --- | --- |
|  | True cases | True controls | TP | FP | PPV | Recall | FPR | TP | FP | PPV | Recall | FPR |
| **T2DM** | 9,293 | 23,754 | 8,978 | 578 | 53.3% | 96.6% | 2.4% | 0 | 0 | - | 0.0% | 0.0% |
| **Dementia** | 2,985 | 77,575 | 729 | 11 | 96.3% | 24.4% | 0.01% | 2,388 | 583 | 71.4% | 80.0% | 7.5% |
| **Hypo-thyroidism** | 2,030 | 25,760 | 2,029 | 258 | 9.6% | 99.9% | 1.0% | 2,029 | 1,065 | 10.7% | 99.9% | 4.1% |

TP=true positive; FP=false positive; FDR=false discovery rate; T2DM=type 2 diabetes mellitus.

The case (and control) prevalences for the eMERGE algorithms in our population were 10.9% (28.0%), 3.5% (91.5%), and 2.4% (30.4%) for T2DM, dementia, and hypothyroidism, respectively. The GPT-4 generated dementia algorithm achieved the highest PPV of 96.3% at the expense of a low recall (24.4%). This result was primarily attributable to the relatively restrictive inclusion criteria, which required medication prescription coupled with the co-occurrence of either diagnosis codes, symptoms, or cognitive assessment tests or procedures. The GPT-4 generated algorithms for T2DM and hypothyroidism, as well as the GPT-3.5 generated algorithm for hypothyroidism, achieved high recall but at the cost of lower PPV. In contrast, the GPT-3.5 generated algorithm for dementia achieved balanced PPV and recall.

## DISCUSSION

EHR phenotyping is a critical area of modern observational clinical research, yet it commonly demands substantial resources. In this study, we explored the effectiveness of LLMs in creating preliminary versions of computable phenotyping algorithms, with the ultimate goal of streamlining the EHR phenotyping process.

Not all LLMs we tested were well-suited for phenotyping. GPT-4 and GPT-3.5 significantly outperformed both Claude 2 and Bard in their ability to generate executable and accurately SQL-formatted phenotyping algorithms. One of the reasons for this discrepancy was Claude 2’s tendency to represent concepts using numerical concept codes, without specifying what clinical criteria these concept codes were intended to capture. Five of the algorithms generated by Bard did not follow the OMOP CDM. Four of these algorithms referenced columns that did not exist in the OMOP CDM. These corresponded to “patient_id” for both prompting strategies of dementia, “observation_fact” and “measurement_fact” for the α-prompting strategies of T2DM, and “occurrence_age” for the β-prompting strategy of T2DM. Additionally, two of the algorithms searched for concepts in the wrong tables. The α-prompting algorithm for dementia attempted to use the “person” table to extract all concepts, including diagnosis codes and medications, which would not be found in this table, whereas the β-prompting algorithm for hypothyroidism looked for signs and symptoms in the Measurement table. We consider these behaviors to be indicative of LLM hallucinations in the context of EHR phenotyping. Given the poor performance of the phenotyping algorithms generated by Claude 2 and Bard, we focused the remaining analysis on phenotyping algorithms generated by GPT-4 and GPT-3.5.

Both GPT models were able to follow instructions and identify relevant concepts for the three selected phenotypes. The phenotyping algorithms generated by these models contained reasonably accurate diagnosis codes, related lab tests, and key medications. The concepts identified largely overlapped with the ones used by domain experts. We found that GPT-4 generally demonstrated slightly superior performance compared to GPT-3.5 by identifying more medications and providing more appropriate thresholds for lab values (**Table 1**). Moreover, the GPT-4 and GPT-3.5 generated algorithms were able to identify additional potentially useful criteria, including a variety of symptoms and clinical signs for each phenotype, which have not usually been incorporated in any eMERGE algorithms.

Despite the merits of GPT-4 and GPT-3.5, they produced phenotyping algorithms containing incorrect criteria. For example, both GPT-4 and GPT-3.5 erroneously considered the ICD-10 code F00 as a relevant diagnosis code for dementia, whereas our prompts specifically required ICD-10-CM codes. Additionally, GPT-3.5 occasionally selected inappropriately broad ICD-9-CM codes, such as 250* for T2DM, inadvertently encompassing diagnoses related to other types of diabetes. Also, both GPT models missed some key ICD codes and medications. For example, while the query correctly identified patients with ICD-10-CM code G30 as dementia cases, they overlooked patients with more specific codes such as G30.0, G30.1, G30.8, and G30.9. The algorithms produced by both GPT models generally generated a shorter list of medications compared to their corresponding eMERGE algorithms, with the exception of the GPT-4 produced algorithm for hypothyroidism. In this case, GPT-4 interpreted the hypothyroidism phenotype more broadly to include not only endogenous causes of hypothyroidism (as in the eMERGE algorithm) but also exogenous causes (not included in the eMERGE algorithm). As a result, the medications specified by the GPT-4 hypothyroidism algorithm included both drugs used for treatment of hypothyroidism and drugs with potential to cause hypothyroidism (i.e., lithium, amiodarone, Lithobid, Cordarone, Nexterone, Pacerone). Moreover, certain thresholds set for lab values were inaccurate, such as “fasting blood glucose ≥6.5” for T2DM.

As noted above, compared to the eMERGE algorithms, both GPT-4 and GPT-3.5 introduced some new concepts, including signs and symptoms such as fatigue, cold intolerance, and weight gain. Many of these concepts are infrequently used by domain experts in algorithm generation due to their low specificity. Including these low-specificity concepts might not substantially enhance the recall of the algorithms and could potentially diminish the algorithm’s PPV. Despite this, they showed promise in identifying noteworthy concepts, such as the Mini-Mental State Examination and Montreal Cognitive Assessment for dementia.

Finally, the evaluated LLMs exhibited immature capability in organizing phenotyping criteria with the proper logic. The SQL queries generated by GPT-4 and GPT-3.5 were predominantly characterized by a single logical operator (AND or OR), resulting in phenotyping algorithms that were either excessively restrictive or overly broad.

Collectively, our findings highlight that LLMs have the potential to produce helpful preliminary phenotyping algorithms by identifying relevant clinical concepts and criteria. As a result, we believe they have the potential to accelerate the creation of EHR-based phenotypes; however, clinical phenotyping expertise, familiarity with EHR data models, and SQL programming skills remain critical for assessing and subsequently enhancing the efficacy of LLM-produced algorithms. At present, LLMs fall short of creating ready-to-use algorithms without revision. Therefore, incorporating a human-in-the-loop approach is necessary at this stage to ensure the algorithms’ practical applicability and accuracy. Still, this preliminary step would allow for a shift from traditional approaches where domain experts independently conduct comprehensive literature review and evidence synthesis—a process that is both time-intensive and challenging—to a more efficient and manageable model, where the domain experts’ role evolves to assessing and refining algorithms generated by LLMs.

## Limitations and future work

Several limitations need to be highlighted as potential opportunities for future improvement. First, this pilot study was limited in scope and did not investigate all the possible tools, options, and capabilities of the LLMs listed here. Additionally, we did not fully explore the improvements, if any, of different prompt-engineering strategies, in-context learning, or fine-tuning. We are reporting initial observations from using the most basic and widely accessible version of these LLMs, i.e., the chat box interface with default settings available through the websites of each of the LLMs. Using an API would allow for selection of additional parameters, which may affect performance. Second, as a pilot study, this research focused on prompting LLMs to generate algorithms for identifying phenotype cases. The capability of LLMs in generating algorithms to identify controls also needs to be evaluated. Third, we tested solely on proprietary models and their default configurations. It is important to assess both proprietary models and the leading open-source models (e.g., Llama 2), especially when they are enhanced with fine-tuning capabilities and knowledge integration. Fourth, our design of prompts did not consider optimizing for execution efficiency of the SQL queries. Consequently, LLMs often produced SQL queries with suboptimal query structures. Fifth, this study only considered phenotypes for three common diseases. Phenotyping rare diseases may present different challenges particularly when there is limited relevant online content for a particular disease. Sixth, there is a chance that the phenotyping algorithms produced by LLMs might include clinical concepts that are not universally adopted by the OMOP databases of all healthcare organizations. As a result, this could lead to uncertainty in the algorithms’ effectiveness in patient identification in such situations. Nevertheless, we believe this issue can be addressed through a human-in-the-loop approach, which involves initially determining the concepts that are either not implemented or included, followed by re-prompting the LLMs to fine-tune the algorithm to exclude these concepts. Seventh, the consistency of responses generated by proprietary LLMs varies over time, warranting consideration for potential future investigations. New LLMs are being rapidly deployed, and our future efforts will involve exploring alternative advanced models, such as ResearchGPT^33^ and Google Gemini^34^. Additionally, we will be delving into more refined control and customization in the generation process through prompt engineering to achieve desired performance levels.

## CONCLUSION

GPT-4 and GPT-3.5 of ChatGPT are capable of producing phenotyping algorithm drafts that align with a standard CDM. These models can reasonably identify relevant clinical inclusion and exclusion criteria that can be used in an initial draft phenotype algorithm. Nevertheless, expertise in informatics and clinical experience is still required to assess and further refine LLM-generated phenotyping algorithms for improving EHR phenotyping accuracy.

## Data Availability

Source code for results and LLM-generated SQL-formatted algorithms are shared at https://github.com/The-Wei-Lab/LLM-Phenotyping-2024.

## AUTHOR CONTRIBUTIONS

C.Y. designed the interactions with the large language models (LLM), collected data, and analyzed the results. W.Q.W. and H.H.O. implemented the algorithms produced by LLMs. M.E.G., V.E.K., and W.Q.W. performed expert review for the produced algorithms. M.E.G., C.Y., H.H.O., V.E.K., and M.S.K. drafted the paper. W.Q.W., B.A.M., W.C.S., A.L.D., Q.P.F., and C.M.S. critically revised the paper. D.M.R. and J.F.P. reviewed the paper and provided important intellectual content. W.Q.W. supervised this study. All authors approved this study.

## CODE AVAILABILITY

Source code for results, LLM-generated SQL-formatted algorithms, and implemented SQL code are shared at https://github.com/The-Wei-Lab/LLM-Phenotyping-2024.

