## Supplementary data for "Large Language Models Facilitate the Generation of Electronic Health Record Phenotyping Algorithms"

**S.1 Prompt design**

**Table 1. Prompts with Type 2 diabetes as an example.**

| **The first-round generation of phenotyping algorithm (with two prompts executed sequentially in the same context)** | **Prompt α1: ask large language models (LLM) to return the phenotyping algorithm of a specific phenotype** |
| --- | --- |
|  | You are an expert in disease phenotyping.    Please provide the phenotyping algorithm for type 2 diabetes.    Let's think step by step:  1. List the critical criteria to consider.  2. Determine how these criteria should be combined.  3. Derive the final algorithm.    Instructions to follow:  1. Use "AND", "OR", and "NOT" to organize the algorithm.  2. Use Observational Medical Outcomes Partnership (OMOP) concepts for diagnoses, symptoms, procedures, laboratory tests, medication, etc.  3. Use ICD9CM and ICD10CM codes for diagnosis concepts.  4. Use both generic names and brand names of medications for medication concepts.  5. Use SQL style to organize the final algorithm. |
|  | **Prompt α2: ask LLMs to return the corresponding SQL query for implementation in an OMOP CDM-based EHR database** |
|  | Please generate the executable SQL query of the algorithm above so that the cohort of type 2 diabetes can be found from an EHR database that follows the OMOP Common Data Model.    Instructions to follow:  1. Use ICD9CM and ICD10CM codes for diagnosis concepts.  2. Use the OMOP concept names for the concepts that are not diagnoses.  3. Use both generic names and brand names of medications for medication concepts.  4. Generate a single SQL query and do not use the WITH clause.  5. Do not create placeholders for any concepts. |
| **The second-round generation of phenotyping algorithm (in a new LLM instance)** | **Prompt β: ask LLMs to analyze the 5 independently produced phenotyping SQL queries and then provide a refined version** |
|  | You are an expert in disease phenotyping.    Below are five SQL queries (in triple quotation marks) for phenotyping type 2 diabetes in an EHR database. Your task is to check the quality of these algorithms and provide a refined version so that the cohort of type 2 diabetes can be accurately found from an EHR database that follows the OMOP Common Data Model.    Let us think step by step.  1. Read each of the five algorithms, summarize the correct criteria, specify the missing criteria, if any, and specify the incorrect criteria, if any.  2. Considering the summarization, generate a refined version of the phenotyping algorithm as the final algorithm.    Instructions to follow when generating the final algorithm:  1. Use ICD9CM and ICD10CM codes for diagnosis concepts.  2. Use the OMOP concept names for the concepts that are not diagnoses.  3. Use both generic names and brand names of medications for medication concepts.  4. Generate a single SQL query and do not use the WITH clause.  5. Do not create placeholders for any concepts.    Query 1: “““”””  Query 2: “““”””  Query 3: “““”””  Query 4: “““”””  Query 5: “““””” |

**S.2 Expert evaluation rubric**

**Table 2. Evaluation rubric for expert review.**

| **Task** | **Axis** | **Question** | **Level** | **Evidence description** |
| --- | --- | --- | --- | --- |
| **1** | **The ability to follow instructions.** | How does the SQL query follow the instructions specified in the prompt (see “file”)? (“No” means a violation.)   1. Whether the generated query follows the OMOP Common Data Model (e.g. whether all of the incorporated table names and column names exist in OMOP CDM)? 2. Whether the generated query uses ICD9CM and/or ICD10CM codes to represent diagnoses? 3. Whether the generated query uses the OMOP concept names for the concepts that are not diagnoses? 4. Whether the generated query provides both the brand names and the generic names of medications, when the medication concepts were mentioned? 5. Whether the LLM generates a single SQL query, rather than separated queries? 6. Whether the generated query creates placeholders for any concepts (e.g., a string indicating that it needs a diagnosis code here or an evident fake ID, say “01234567”)? | Good (0-2 violations) | (Please provide evidence briefly as the justification of the provided score) |
|  |  |  | Medium (3-4 violations) |  |
|  |  |  | Poor (5-6 violations) |  |
| **2** | **The ability to generate reasonably good phenotyping algorithms.** | Whether the generated query incorporates relevant phenotyping criteria, e.g. ICD codes, medications etc.? Consider the following three dimensions.   1. Does the query miss important criteria? 2. Does the query misidentify criteria? 3. Does the query identify meaningfully novel criteria that are relevant? | Good (having no problems or only minor problems) | (Please provide justifications) |
|  |  |  | Medium (having some problems) |  |
|  |  |  | Poor (having major problems) |  |
| **3** | **The ability to generate executable SQL code.** | How much manual effort is required to finetune the generated SQL code so that it can run in EHR databases that follow the OMOP Common Data Model? (Please focus on evaluating the necessary level of finetuning, rather than the degree to which the query should be optimized.) | Good (minor) | (Please provide justifications) |
|  |  |  | Medium (moderate) |  |
|  |  |  | Poor (major) |  |

**S.3 Implementation of algorithms**

After expert assessment, it was decided to implement the phenotype algorithms generated by GPT-4 and GPT-3.5 in the β-prompting scenario as they had the highest rated executions scores (2.4 and 2.3 out of 3, requiring minimal/moderate modification). Our implementation experience generally matched this initial assessment. Overall observations were that the SQL algorithms were well-formatted with comments and clear to read and understand.

Implementation required technical knowledge of the OMOP CDM and SQL programming language to modify LLM-generated algorithms so that they can be successfully executed in VUMC EHR database, including removal of extraneous/incorrect characters, sourcing appropriate tables, and modifying operations that combine tables. To ensure evaluation validity, no clinical domain knowledge of phenotyping was used. The list of edits to the LLM-generated algorithms were as follows:

1. Include or edit the VUMC-specific database name where the OMOP tables are stored.
2. Remove extraneous texts like “ICD9CM:” that appear in front of quoted concepts.
3. Replace incorrect wildcard characters.
4. Replace searching for concepts in the Concept table with searching on standardized clinical data tables containing the names of those concepts, e.g. search for ICD codes in the condition_occurence table in the condition_source_value column.

5. Replace searching for exact concept names for drugs, laboratory tests, and symptoms using an “IN” statement with searching for additional variations of names using a “LIKE” operator with wildcards at the beginning and end of the search terms and removing case-sensitivity with the “LOWER()” function.
6. Replace “OR” with “UNION” for algorithmic efficiency when searching for concepts across multiple tables.
7. Replace consecutive “LEFT JOIN” statements that combine multiple tables using “OR” statements, with a “UNION” operation on the tables that have been left joined.

Edits 1, 2 and 3 were clarification edits. In Edits 4 and 5, it should be noted that LLMs typically reply on exact matches of name strings when it comes to symptoms, drugs, and laboratory tests. This potentially overlooked a non-negligible number of name variations in EHR databases. Moreover, LLM-produced algorithms searched for concepts either via the Concept table or through source value columns in other tables (such as “condition_occurrence”, “procedure_occurrence”, “measurement”, “drug_exposure” and “observation”). Our observations have shown that the latter method tends to yield a higher number of results. This occurs because not all name variations of clinical concepts are allocated a concept ID in the Concept table. Edits 6 and 7 were made in order for the algorithms to execute in an appropriate amount of time without changing the results.

Since all the required edits were limited to technical knowledge regarding OMOP and SQL programming language, we expect that the limited manual work needed to realize them can be further reduced or eliminated with appropriate prompt-engineering, in-context learning, or fine-tuning strategies in the future.

Modifications made to each algorithm for implementation purposes are summarized in Table 3. The versions of the Phenotyping algorithms before and after modifications are available at <https://github.com/The-Wei-Lab/LLM-Phenotyping-2024> .

**Table 3. Modifications made for algorithm implementation.**

|  | **Phenotype** | | | | | |
| --- | --- | --- | --- | --- | --- | --- |
|  | **T2DM** | | **Dementia** | | **Hypothyroidism** | |
| **Edits**^a^ | **GPT-4** | **GPT-3.5** | **GPT-4** | **GPT-3.5** | **GPT-4** | **GPT-3.5** |
| **Edit 1:**    **Include/edit database names** | Add site-specific database name in front of OMOP tables | | | | | |
| **Edit 2:**    **Remove extraneous texts appear in front of concepts** | - | - | Remove prefixes “ICD9CM:”, “ICD10CM:”, and “OMOP” | - | - | - |
| **Edit 3:**    **Replace incorrect wildcard characters** | Change wildcard character from ‘x’ to ‘%’ | - | - | - | - | - |
| **Edit 4:**    **Change concept search strategy from concept table to standardized clinical data tables** | Search concepts in the source value columns of the standardized clinical data tables | Search concepts in the source value columns of the standardized clinical data tables | - ^b^ | Search concepts in the source value columns of the standardized clinical data tables | Search concepts in the source value columns of the standardized clinical data tables | Search concepts in the source value columns of the standardized clinical data tables |
| **Edit 5:**    **Expand pattern searching to capture concept name variations** | Add “LOWER()” and “LIKE” operators with wildcards for concept names | | | | | |
| **Edit 6:**    **Improve algorithmic efficiency** | - | - | - | Replace “OR” with “UNION” as searching for concepts across multiple tables | - | Replace “OR” with “UNION” as searching for concepts across multiple tables |
| **Edit 7:**    **Replace consecutive left join to accelerate execution** | Replace consecutive left joins that combine multiple tables using “OR” statements in a “WHERE” clause, with a union of joined tables | Replace consecutive left joins that combine multiple tables using “OR” statements in a “WHERE” clause, with a union of joined tables | Replace consecutive left joins that combine multiple tables using “OR” statements in a “WHERE” clause, with a union of joined tables | - | Replace consecutive left joins that combine multiple tables using “OR” statements in a “WHERE” clause, with a union of joined tables | - |

^a^ Edits are further explained in Supplement S.3 above.

^b^ The LLM suggested using concept IDs in standard clinical tables, but only provided concept names. We modified this by using source value columns instead.

**S.4 The comparison of clinical concepts**

**Table 4. A comparative summary of medical concepts utilized in GPT-4 (β), GPT-3.5 (β), and eMERGE phenotyping algorithms.** For each phenotype, we show the concepts that all three algorithms identified and the concepts that were unique to each algorithm (relative to the eMERGE algorithm). The underlined concepts indicate concepts unique to GPT-4 or GPT-3.5.

|  | **Type 2 diabetes mellitus** | | | |
| --- | --- | --- | --- | --- |
|  | **Common concepts** | **Unique concepts** | | |
|  |  | **eMERGE** | **GPT-4 (**β**)** | **GPT-3.5 (**β**)** |
| **Diagnoses** | 250.*0, 250.*2, E11.* | O24.11, E11 | n/a | Includes 250*, which is inaccurate (encompasses codes for type 1 diabetes mellitus) |
| **Lab tests or procedures** | Hemoglobin A1c >6.5, Fasting blood glucose | Hemoglobin A1c ≥6.5, Fasting blood glucose ≥125, Random glucose >200 | Hemoglobin A1c >6.5, Fasting blood glucose >126, Oral glucose tolerance test >200 | Hemoglobin A1c ≥6.5, Fasting blood glucose ≥6.5  (Threshold for fasting blood glucose is inaccurate)  BMI ≥25 |
| **Medications** | Metformin, Glipizide | Acetohexamide, Dymelor, Tolazamide, Tolinase, Chlorpropamide, Diabinese, Glucotrol, Glipizide, Glucotrol XL, Glyburide, Micronase, Glynase, Diabeta, Glimepiride, Amaryl, Repaglinide, Prandin, Nateglinide, Starlix, Glucophage, Rosiglitazone, Avandia, Pioglitazone, Actos, Troglitazone, Rezulin, Acarbose, Precose, Miglitol, Glyset, Sitagliptin, Januvia, Exenatide, Byetta | Glucophage, Glucotrol, Glyburide, Diabeta, Glynase, Glucophage XR, Glucotrol XL | Sitagliptin |
| **Symptoms or signs** | n/a | n/a | Polyuria, Polydipsia, Unexplained weight loss | Polyuria, Polydipsia, Unexplained weight loss  (Looking specifically for absence of these symptoms) |
| **Exclusion criteria** | n/a | 250.*1, 250.*3, 250.10, 250.12, E10, E08.42, E13.42, 357.2, 362.0 | 250.*1, 250.*3, E10.* | n/a |
|  | **Dementia** | | | |
|  | **Overlapping concepts** | **Specific concepts** | | |
|  |  | **eMERGE** | **GPT-4 (**β**)** | **GPT-3.5 (**β**)** |
| **Diagnoses** | n/a | 290.0, 290.10, 290.11, 290.12, 290.13, 290.20, 290.21, 290.3, 290.40, 290.41, 290.42, 290.43, 291.0, 291.1, 291.2, 292.82, 294.8, 294.10, 294.11, 331.0, 331.11, 331.19, 331.82, F01, F01.5, F01.50, F01.51, F02, F02.8, F02.80, F02.81, F03, F03.9, F03.90, F03.91, G30, G30.0, G30.1, G30.8, G30.9, G31.0, G31.01, G31.09, G31.1, G31.83 | 290, 294.1, 331.0, G30, F00, F03  (F00 is an ICD-10 code, not an ICD-10-CM code) | 290*, F00, F01  (F00 is an ICD-10 code, not an ICD-10-CM code) |
| **Lab tests or procedures** | n/a | n/a | Mini-Mental State Examination (MMSE), Montreal Cognitive Assessment (MoCA), Neuropsychological testing, Brain imaging procedure  (No value thresholds or other specifications applied to test results) | Neuropsychological testing, MRI scan, CT scan  (No value thresholds or other specifications applied to test results) |
| **Medications** | Donepezil, Aricept, Memantine, Namenda | Galantamine, Razadyne, Reminyl, Nivalin, Rivastigmine, Exelon, Tacrine, Cognex, Axura, Ebixa, Akatinol | Rivastigmine, Exelon, Galantamine, Razadyne | n/a |
| **Symptoms or signs** | n/a | n/a | Memory loss, Impaired reasoning, Impaired communication | Memory loss, Cognitive impairment, Disorientation, Language problems, Behavioral changes |
| **Exclusion criteria** | n/a | n/a | Thyroid function test, B12 level, Syphilis test  (No value thresholds or other specifications applied to test results) | n/a |
|  | **Hypothyroidism** | | | |
|  | **Overlapping concepts** |  | | |
|  |  | **eMERGE** | **GPT-4 (**β**)** | **GPT-3.5 (**β**)** |
| **Diagnoses** | 244.9, E03.8, E03.9 | 244, 244.8, 245, 245.2, 245.8, 245.9, E00, E00.0, E00.1, E00.2, E00.9, E01.8, E02, E03.0, E03.1, E03.2, E03.3, E89.0 | 244.0, 244.1, 244.2, 244.3, 244.8, E02, E89.0 | E03.0, E03.1, E03.2, E03.3, E03.5, E03.89 |
| **Lab tests or procedures** | Thyroid-stimulating hormone (TSH) | TSH >5, Free T4 <0.5, Anti-thyroglobulin antibodies (H-TGA, ThyrAB, AThyg- positive), Anti-thyroperoxidase (H-TPO, TPO, AThyP-positive), Anti-thyroid antibodies (ThyAb-positive) | Serum TSH >high range,  Serum free T4 <low range | TSH >4.5 |
| **Medications** | Levothyroxine, Synthroid | Levoxyl unithroid, Armour thyroid, Desiccated thyroid, Cytomel, Triostat, Liothyronine, Synthetic triiodothyronine, Liotrix, Thyrolar | Levoxyl, Euthyrox, Eltroxin, Liothyronine, Cytomel, Desiccated thyroid, Armour thyroid, Nature-Throid, WP Thyroid, Lithium, Amiodarone, Lithobid, Cordarone, Nexterone, Pacerone | n/a |
| **Symptoms or signs** | n/a | n/a | Fatigue, Weight gain, Cold intolerance, Dry skin, Bradycardia, Hair loss | Fatigue, Cold intolerance, Weight gain, Dry skin, Constipation, Bradycardia |
| **Exclusion criteria** | n/a | 193*, 242, 242.1, 242.2, 242.3, 242.9, 244, 244.1, 244.2, 244.3, 258.*,…  (Long list of exclusion codes, including CPT codes) | Thyroid surgery, Radioactive iodine treatment | n/a |
